# Synaptic density (SV2A) tracks illness chronicity rather than symptom severity in major depressive disorder

**DOI:** 10.64898/2026.10.27.26366602

**Authors:** Hampus Yngwe, Pontus Plavén-Sigray, Max Andersson, Martin Bolin, Maria Beckman, Carl-Johan Ekman, Mikael Tiger, Johan Lundberg

## Abstract

Synaptic loss has been proposed as a key mechanism in major depressive disorder (MDD). Positron emission tomography with [¹¹C]UCB-J enables in vivo quantification of synaptic vesicle glycoprotein 2A (SV2A), a proxy marker of synaptic density. A landmark study by Holmes et al reported an inverse association between depression severity and SV2A binding in frontolimbic regions. As a replication, we examined this association in [¹¹C]UCB-J PET data from 34 unmedicated individuals with MDD, focusing on the dorsolateral prefrontal cortex, anterior cingulate cortex, and hippocampus. We found no support of an association between MADRS scores and SV2A binding in the primary regions of interest (r = 0.18–0.26, all p ≥ 0.15). Findings were similar in unadjusted and age- and sex-adjusted models. However, adjustment for duration of the current depressive episode and lifetime depressive episodes yielded positive associations across primary regions. In post hoc analyses, longer current episode duration, but not lifetime depressive episodes, was negatively associated with SV2A binding across primary regions. Thus, our findings did not support an association between depression severity and synaptic density, contrary to a previous report; instead, the exploratory findings suggest that SV2A binding may be related to episode duration rather than current symptom burden.

## Introduction

The synaptic loss hypothesis of major depressive disorder (MDD) posits that reduced synaptic connectivity may disrupt signalling in affective circuits, thereby contributing to depressive symptoms^1,2^. This hypothesis is supported by animal research and post-mortem studies demonstrating reduced synapse number and alterations in synapse-related genes in depression^1,3–5^. In contrast, broader plasticity i.e. metaplasticity frameworks emphasize impaired capacity for adaptive synaptic remodeling in depression, suggesting that loss of synapses may not capture the full pathology^4,6^.

Positron emission tomography (PET) radioligand [^11^C]UCB-J, targeting synaptic vesicle glycoprotein 2A (SV2A), selectively expressed in presynaptic boutons, has been proposed as a proxy marker of synaptic density^7,8^. This has enabled in vivo investigation of synaptic alterations in neuropsychiatric conditions in which synaptic pathology has been implicated ^9–11^

In a seminal paper, Holmes et al reported a negative association between depressive symptom severity and SV2A binding in frontolimbic regions in a sample including MDD, Post-Traumatic Stress Disorder (PTSD) and comorbid MDD/PTSD^12^. Individuals with more accentuated depressive symptoms also showed lower SV2A binding compared to healthy controls. Consistent with these clinical findings, preclinical studies have also reported reduced SV2A binding in stress-based depression models in rodents^13,14^. Together, these findings indicate that MDD may be associated with synaptopathy^15^ and raise the question of whether successful treatment might rescue synapse number and function.

A subsequent PET–MRI study, albeit in late-life depression, found no association between depressive symptom severity and SV2A binding^16^. Several investigations have used radiolabelled UCB-J to track treatment-related synaptic changes after antidepressants and psychedelic interventions. However, although Raval et al reported increased [^3^H] UCB-J binding following psilocybin administration in pigs, robust evidence from human PET studies that SV2A binding is responsive to treatment effect is limited. Treatment with ketamine and ECT did not yield increased SV2A binding at the group-level, despite substantial clinical improvement, although exploratory analyses suggested increases in patients with a baseline SV2A deficit following ketamine^17^ and in patients achieving remission after ECT^18^.

Similarly, although escitalopram administration in healthy volunteers did not produce a primary group-level increase, exploratory analyses suggested increases as a function of treatment duration^19^.

It is not established whether synaptic alterations in MDD represent pre-existing vulnerability, causal mechanisms or consequences of prolonged or recurrent depressive episodes. A complementary perspective comes from the structural magnetic resonance imaging (MRI) literature, which has linked hippocampal volume reductions - one of the most replicated in vivo structural findings in MDD - with illness chronicity^20–23^. Mechanistically, this is consistent with findings in animal models showing that dendritic atrophy in the hippocampus and prefrontal cortex emerge after weeks of sustained stress exposure.^24–27^ Grey matter reductions in depression have been proposed to largely reflect dendritic and synaptic loss^28^; however, prior SV2A PET work has not shown a corresponding association between SV2A binding and volumetric alterations^9,16^. This suggests that grey matter volume and SV2A binding may capture partly distinct aspects of disease biology, providing a rationale for examining whether SV2A binding may be associated with disease course in MDD.

To summarize, the lack of robust treatment-related changes in SV2A binding, and its limited association with short-term symptom variation, has raised questions about the suitability of [¹¹C]UCB-J as a biomarker for MDD. Given this background, we wanted to revisit the original study to see whether the findings reported by Holmes et al. could be replicated. Because the original sample was transdiagnostic, combining MDD, PTSD, and comorbid cases, the present cohort allowed us to test whether the association generalizes to a MDD sample.

In this conceptual replication, we hypothesized that higher depression severity scores would be associated with lower distribution volume (*VT*) of [¹¹C]UCB-J in the dorsolateral prefrontal cortex, anterior cingulate cortex, and hippocampus. In exploratory analyses, we additionally examined whether chronicity-related clinical measures, including lifetime depressive episodes and duration of the current depressive episode, were associated with SV2A binding across the same regions.

## Methods

### Participants

The sample comprised 34 men and women aged 20-65 years with an episode of recurrent major depressive disorder and without ongoing treatment. Participants were recruited on social media and Google Ads. The current depressive episode was required to have lasted at least 30 days but less than five years, and to be in the moderate-to-severe range, defined as a MADRS score ≥22 at screening. Exclusion criteria were bipolar and psychotic disorders, substance abuse disorder and suicidality, as well as diverse somatic disorders (complete inclusion/exclusion criteria found on ClinicalTrials.gov; study ID NCT04630964). All participants provided written informed consent. One additional participant had been recruited but later withdrew consent and was therefore not included in the present analyses. Seven participants tapered medication before baseline PET, with a washout period of at least five elimination half-lives; medication details are provided in Supplementary Table 7. The protocol was approved by the Swedish Ethical Review Authority and the Swedish Medical Products Agency. The study was conducted in accordance with the Declaration of Helsinki.

### Assessments

This study used baseline symptom assessments from the PSIPET clinical trial. Symptom severity was assessed using clinician-rated Montgomery–Åsberg Depression Rating Scale (MADRS), in contrast to the primary study, which used HAMD-17.

At study inclusion, chronicity-related variables were assessed through participant reports during the clinical interview. Participants were asked to estimate the number of distinct depressive episodes they had experienced during their lifetime, and the duration of the current depressive episode, recorded in months.

### Magnetic Resonance Imaging

A structural MRI scan was performed 7–35 days prior to dosing at the Solna MRI Centre, Department of Clinical Neuroscience, Karolinska Institutet. The first 25 participants were scanned on a 3-Tesla General Electric Discovery MR750 scanner and the remaining 9 participants on a 3-Tesla GE SIGNA Premier scanner.

### Positron Emission Tomography

Participants were examined using [¹¹C]UCB-J PET on a High-Resolution Research Tomograph (HRRT; Siemens Molecular Imaging). Participants were positioned supine in the scanner with the head fixated to minimize motion.

[¹¹C]UCB-J was synthesized at the Karolinska PET Centre in accordance with Good Manufacturing Practice. The radiotracer was administered as an intravenous bolus (∼400 MBq ±10%), followed by a saline flush.

A transmission scan was acquired for attenuation correction, followed by a 90-minute dynamic emission scan acquired in list mode. Dynamic images were reconstructed into 37 frames (8 × 15 s, 8 × 30 s, 4 × 60 s, 5 × 120 s, 10 × 300 s, and 2 × 600 s) using OP-3D-OSEM with point spread function modelling.

Arterial blood sampling was performed via a radial artery cannula for derivation of the metabolite-corrected arterial input function. Continuous sampling was performed during the initial phase, followed by discrete manual sampling. The parent fraction in plasma and radiometabolite profile were determined using protein precipitation followed by reversed-phase radio-HPLC. Head motion was estimated using frame-to-first-frame realignment in SPM. For scans with translational displacement >4 mm, motion correction was performed by frame-wise coregistration to the corresponding transmission image.

Structural T1-weighted MRI scans were segmented and parcellated into regions of interest (ROIs) using FreeSurfer based on the Desikan–Killiany atlas (cortical ROIs) and FreeSurfer subcortical segmentations for hippocampus. PET images were coregistered to individual MRI scans, and regional time-activity curves were extracted for all ROIs. No correction for partial-volume effects was applied.

Kinetic modelling was performed using the open-source R package *kinfitr*^29^.

In line with Holmes et al., the primary outcome measure was total distribution volume (*VT*), estimated over the 90-min acquisition using a one-tissue compartment model with a metabolite-corrected arterial input function and blood volume fraction fixed to zero.

### Statistical analysis

The present substudy was conducted within the framework of the PSIPET clinical trial.

All analyses were conducted in R (version 4.1.1). The aim was to perform a conceptual, rather than a direct replication^30,31^ of the association between depressive symptom severity and synaptic density indexed by SV2A binding, reported by Holmes et al.

Primary correlation and regression analyses included 32 participants with complete MADRS and *VT* data; *VT* could not be estimated for two participants because arterial input-function data were unavailable. Correlations involving current episode duration included 31 participants with available episode-duration and *VT* data, while fully adjusted models included 28 participants with complete data on all model variables.

Pearson correlation analyses were used to assess the association between depressive symptom severity (MADRS) and [¹¹C]UCB-J total distribution volume (*VT*) in three a priori regions of interest: anterior cingulate cortex, hippocampus, and dorsolateral prefrontal cortex^12^. Corresponding 95% confidence intervals were calculated.

Linear regression models were fitted to further examine these associations. Three models were specified: (1) unadjusted; (2) adjusted for age and sex; and (3) additionally adjusted for lifetime number of depressive episodes and estimated duration of the current depressive episode.

Because duration of the current depressive episode was right-skewed, sensitivity analyses were performed using Spearman rank correlations and log-transformed episode duration. Influence diagnostics based on Cook’s distance and leave-one-out analyses were additionally used to assess whether associations were driven by individual observations.

Exploratory analyses were conducted across additional brain regions, including the amygdala, bed nucleus of the stria terminalis, caudate nucleus, centrum semiovale, frontal cortex, gray matter, insula, putamen, thalamus, caudal anterior cingulate cortex, medial orbitofrontal cortex, and rostral anterior cingulate cortex. Associations between MADRS and *VT* for these exploratory regions were conducted identically to the primary regions.

All statistical tests were two-sided with a significance threshold of p < 0.05. For analyses across the three a priori defined primary regions of interest, p-values were adjusted using Bonferroni correction. For exploratory analyses across additional regions, p-values were corrected for multiple comparisons using the Benjamini–Hochberg false discovery rate.

## Results

In the primary regions of interest, MADRS was not significantly associated with [¹¹C]UCB-J *VT*. All correlations were positive in direction, and the 95% confidence intervals excluded negative associations of the magnitude reported by Holmes et al: ACC, r = 0.26, 95% CI [−0.10, 0.56], p = 0.15; hippocampus, r = 0.26, 95% CI [−0.10, 0.56], p = 0.15; and dlPFC, r = 0.18, 95% CI [−0.18, 0.50], p = 0.33 (Supplementary Table 1; Figure 1).

**Figure 1.**
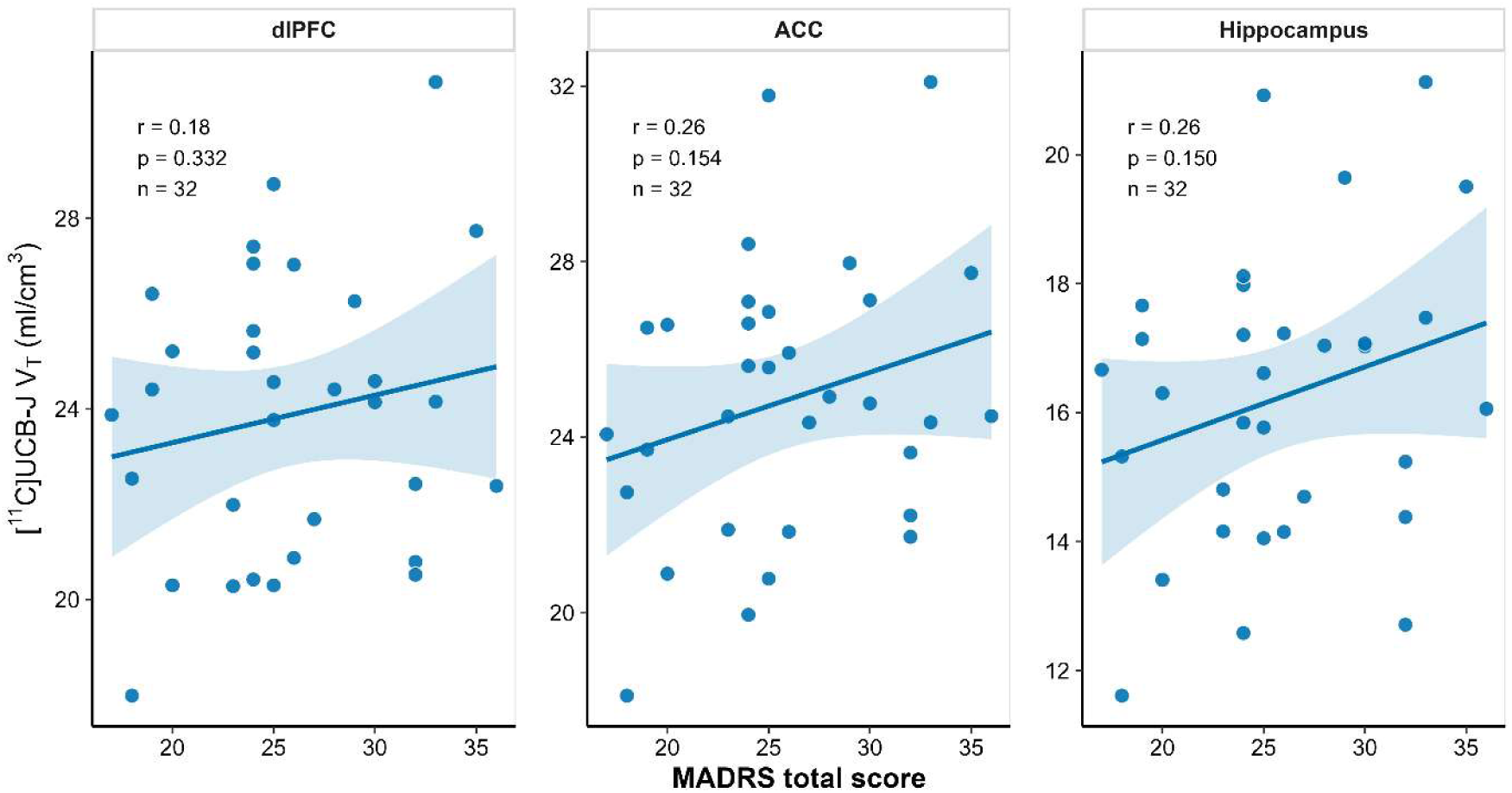
Replication analysis of the association between depressive symptom severity and synaptic density in primary regions of interest. Scatter plots show the relationship between depressive symptom severity (MADRS total score) and synaptic vesicle glycoprotein 2A (SV2A) binding measured with [¹¹C]UCB-J PET (Vₜ, ml/cm³) in the three primary regions of interest examined in the replication analysis: hippocampus, anterior cingulate cortex, and dorsolateral prefrontal cortex. Each point represents an individual participant. Solid lines indicate linear regression fits with 95% confidence intervals. Pearson correlation coefficients (r), corresponding p-values, and sample sizes are shown within each panel.

**Figure 2.**
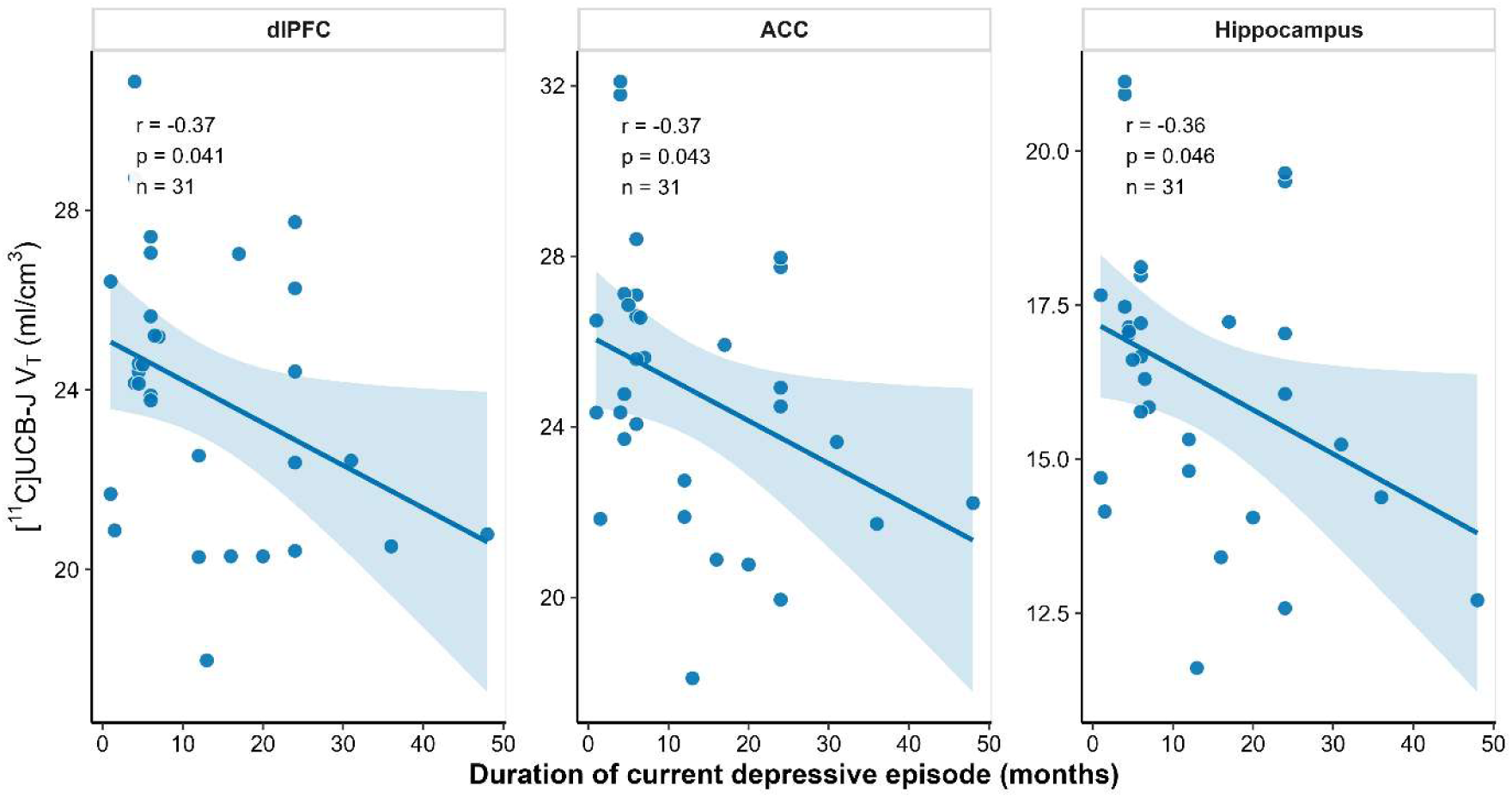
Association between duration of the current depressive episode and synaptic density in primary regions of interest. Scatter plots show the relationship between estimated duration of the current depressive episode, measured in months, and synaptic vesicle glycoprotein 2A (SV2A) binding measured with [¹¹C]UCB-J PET (Vₜ, ml/cm³) in the three primary regions of interest examined in the exploratory analysis: hippocampus, anterior cingulate cortex, and dorsolateral prefrontal cortex. Each point represents an individual participant. Solid lines indicate linear regression fits with 95% confidence intervals. Pearson correlation coefficients (r), corresponding p-values, and sample sizes are shown within each panel.

**Table 1.** Clinical and imaging characteristics of the study sample.

|  | PSIPET sample (N = 34) | Holmes sample (N = 26) |
| --- | --- | --- |
| **Demographics** |  |  |
| Age, mean (SD) | 41.2 (10.2) | 38.92 (12.25) |
| Female sex, n (%) | 20 (58.8) | 10 (38.5) |
| **Clinical Severity and Illness Course** |  |  |
| MADRS total score, mean (SD) | 26.0 (5.1) | 16.28 (7.62) * |
| Duration of current depressive episode (months), median (IQR) † | 6.8 (4.5–21.0) | Not reported |
| Lifetime depressive episodes (number), median (IQR) ‡ | 5.0 (4.0–8.0) | Not reported |
| Duration of illness, years, mean (SD) | Not reported | 18.16 (11.9) |
| **Diagnostic composition, n (%)** |  |  |
| MDD diagnosis | 34 (100) | 21 (80.8) |
| MDD with comorbid PTSD | 2 (5.9) | 10 (38.5) |
| PTSD without MDD | 0 (0) | 5 (19.2) |
| Any comorbid anxiety disorder | 14 (41.2) | Not reported |
| **Treatment and imaging characteristics** |  |  |
| Tapered medication §, n (%) | 7 (20.6) |  |
| [<sup>11</sup>C]UCB-J $V_T$ dlPFC, mean (SD) | 23.87 (2.95) | |
| [<sup>11</sup>C]UCB-J $V_T$ ACC, mean (SD) | 24.83 (3.12) | |
| [<sup>11</sup>C]UCB-J $V_T$ hippocampus, mean (SD) | 16.23 (2.29) | |

Abbreviations: $V_T$ , total distribution volume; MADRS, Montgomery–Åsberg Depression Rating Scale; dlPFC, dorsolateral prefrontal cortex; ACC, anterior cingulate cortex.
Note: The number of available observations varied across variables because of missing data.
\*Holmes et al. used HAMD-17 as the primary severity measure but also reported MADRS scores; the values shown here were calculated from the reported low- and high-severity subgroups.
†Available for 32 participants in the PSIPET sample.
‡Available for 29 participants in the PSIPET sample.
§ Details of medications tapered before baseline PET, including doses, are provided in Supplementary Table 7.

Unadjusted and age- and sex-adjusted regression models similarly showed no significant associations between MADRS and *VT* in the primary regions. In contrast, after additional adjustment for lifetime depressive episodes and duration of the current depressive episode, positive associations emerged in the ACC (b = 0.306, 95% CI [0.082, 0.530], p = 0.010), hippocampus (b = 0.231, 95% CI [0.077, 0.385], p = 0.005), and dlPFC (b = 0.255, 95% CI [0.040, 0.470], p = 0.022), with the ACC and hippocampus remaining significant after Bonferroni correction (Supplementary Tables 2–3).

Exploratory analyses across additional regions showed no significant associations between MADRS and *Vt* in unadjusted models, whereas fully adjusted models revealed positive associations in several regions, including the amygdala and insula (Supplementary Tables 4– 5). Duration of the current depressive episode was negatively correlated with *V*T across all three primary regions (r = −0.36 to −0.37, p = 0.041–0.046). In the fully adjusted models, episode duration remained negatively associated with *Vt* in the ACC (b = −0.171, 95% CI [−0.283, −0.059], p = 0.004), hippocampus (b = −0.121, 95% CI [−0.198, −0.044], p = 0.004), and dlPFC (b = −0.164, 95% CI [−0.272, −0.057], p = 0.004), with all three associations remaining significant after Bonferroni correction. Lifetime number of depressive episodes was not associated with *V_T_*.

Sensitivity analyses supported the robustness of this association. Spearman correlations were similar in direction and magnitude to the Pearson correlations, and episode duration remained negatively associated with *VT* in all three primary regions when log-transformed in the fully adjusted models (all p ≤ 0.007). Although influence diagnostics identified potentially influential observations, the episode-duration coefficient remained negative and statistically significant in all leave-one-out iterations across the three primary regions (Supplementary Table 6).

## Discussion

We did not replicate the association between SV2A binding and depression severity, reported by Holmes et al. in the primary ROIs (ACC, dlPFC, and hippocampus) with point estimates in the opposite direction to the original study. Exploratory analyses, however, suggested that longer current episode duration was associated with lower SV2A binding.

To our knowledge, ours is the first replication study in an MDD sample of comparable age to the original study, although a previous study did not find evidence of an association in late-life depression, in which age-related and neurodegenerative processes may additionally have influenced SV2A binding^16^. The transdiagnostic approach of the original study raises the possibility that the reported association may be specific to a trauma-exposed clinical population, rather than to MDD more broadly. Although alterations in mGluR5 availability, a marker linked to synaptic plasticity, have been reported in PTSD^32^, SV2A-PET studies in PTSD without comorbid depression that could further elucidate this association are still lacking.

An important exploratory finding of the present study was the association between episode duration and SV2A binding across all three primary regions. This finding is consistent with a synaptic loss framework of MDD, in which synapses may be progressively lost in the depressive state, although the cross-sectional design precludes causal inference. Alternatively, lower SV2A binding could represent vulnerability or a negative prognostic factor. The finding aligns with neuroplasticity models of depression, with preclinical findings indicating that stress-related synaptic alterations evolve over weeks^24^ and structural MRI studies linking chronicity-related factors to regional volumetric reductions^20,22,23^. Our study thus extends previous chronicity-related findings based on brain volume to an in vivo marker of synaptic density.

Episode duration and current symptom severity capture distinct aspects of the depressive condition. Symptom rating scales provide limited information about disease course, and severity scores may vary within the same individual and episode^33,34^. Thus, if synaptic loss develops with time spent in a depressive state, cross-sectional severity may be less well suited to index this process. Moreover, while rapid increases in SV2A binding have been observed following psilocybin in animal research^35^, human treatment studies have not demonstrated consistent increases in SV2A binding despite clinical improvement in MDD^17,18^, suggesting that the temporal dynamics of synaptic plasticity may not translate directly from experimental models to human depression. Accordingly, although synaptic density changes may be part of the pathophysiology of depression, their reversal may not mechanistically mediate the antidepressant effect but may occur on a slower timescale as a downstream consequence of the biological and behavioral changes associated with recovery. Comparisons with healthy controls will be important to establish whether lower SV2A binding is associated with MDD, whereas longitudinal imaging studies with repeated PET assessments could determine whether individuals undergo changes in synaptic density across different phases of illness and remission. If the present findings are replicated, framed in terms of clinical staging models of psychiatric illness, this would position SV2A less as a state marker of current depressive symptoms and more as a marker of illness stage or chronicity.

Several limitations should be considered. First, depressive symptom severity was assessed using the MADRS rather than the HAMD-17, reducing direct comparability with the original study. Second, ROI definitions differed: Holmes et al used Automated Anatomical Labeling atlas-derived ROIs, whereas the present study used FreeSurfer-derived parcellations and segmentations. In addition, the original study applied partial-volume correction, which was not applied in the present study. These methodological differences support interpreting the present analysis as a conceptual rather than direct replication. Third, no healthy control group was included, limiting the analyses to within-group associations. Fourth, the parent PSIPET trial was not primarily designed or powered to address the specific research questions examined here, and the sample size was modest, with 32 participants contributing to the primary analyses and 28 to the fully adjusted models, although the total sample was larger than in the Holmes study. Fifth, episode duration was estimated retrospectively and is subject to recall uncertainty. Register-based data on treatment contacts and prescriptions, available in countries such as Sweden, could provide an objective validation of self-reported chronicity in future work. The association was not examined in episodes longer than five years due to the study’s exclusion criteria. Finally, the biological interpretation of SV2A binding remains limited.

## Conclusion

We could not replicate the previously reported association between depression severity and SV2A *VT* in our MDD sample^12^, with correlations in all three primary ROIs non-significant and in the opposite direction to those reported by Holmes et al. Differences in diagnostic composition, including the transdiagnostic nature of the original sample may have contributed to this discrepancy. In contrast, exploratory analyses suggested an association between lower SV2A binding and longer episode duration, rather than current symptom burden. Larger, well-characterized cohorts are needed to clarify the association between symptom severity and SV2A binding. Comparisons with healthy controls and longitudinal studies will be important to determine whether SV2A binding is associated with depression and changes over the course of illness and remission.

## Data Availability

Some data may be made available upon reasonable request, subject to applicable ethical, legal, and data protection requirements.

## Acknowledgments

We thank the study participants, the members of the PET-research group at Karolinska Institutet for their valuable discussions and suggestions on this manuscript. We thank Aleksander Lebedev, Kasim Acar and Predrag Petrovic at Karolinska Institutet for their contributions to collecting and sharing MRI-data used in this project.

## References

1. Duman RS, Aghajanian GK. Synaptic Dysfunction in Depression: Potential Therapeutic Targets. Science. 2012;338(6103):68–72. doi:10.1126/science.1222939

2. Kaiser RH, Andrews-Hanna JR, Wager TD, Pizzagalli DA. Large-scale network dysfunction in Major Depressive Disorder: Meta-analysis of resting-state functional connectivity. JAMA Psychiatry. 2015;72(6):603–611. doi:10.1001/jamapsychiatry.2015.0071

3. Kang HJ, Voleti B, Hajszan T, et al. Decreased expression of synapse-related genes and loss of synapses in major depressive disorder. Nat Med. 2012;18(9):1413–1417. doi:10.1038/nm.2886

4. Castrén E. Neuronal Network Plasticity and Recovery From Depression. JAMA Psychiatry. 2013;70(9):983. doi:10.1001/jamapsychiatry.2013.1

5. Moda-Sava RN, Murdock MH, Parekh PK, et al. Sustained rescue of prefrontal circuit dysfunction by antidepressant-induced spine formation. Science. 2019;364(6436):eaat8078. doi:10.1126/science.aat8078

6. Castrén E. Neurochemistry of the antidepressant effect – a misunderstanding? Eur Neuropsychopharmacol. 2023;75:37–38. doi:10.1016/j.euroneuro.2023.07.001

7. Finnema SJ, Nabulsi NB, Mercier J, et al. Kinetic evaluation and test–retest reproducibility of [11C]UCB-J, a novel radioligand for positron emission tomography imaging of synaptic vesicle glycoprotein 2A in humans. J Cereb Blood Flow Metab. 2018;38(11):2041–2052. doi:10.1177/0271678X17724947

8. Nabulsi NB, Mercier J, Holden D, et al. Synthesis and Preclinical Evaluation of 11C-UCB-J as a PET Tracer for Imaging the Synaptic Vesicle Glycoprotein 2A in the Brain. J Nucl Med. 2016;57(5):777–784. doi:10.2967/jnumed.115.168179

9. Onwordi E et al. Synaptic loss in schizophrenia measured with SV2A PET. Nat Commun. Published online 2020. doi:10.1038/s41467-019-14122-0

10. Chen M et al. Reduced synaptic density in Alzheimer’s disease measured with [11C]UCB-J PET. JAMA Neurol. Published online 2018. doi:10.1001/jamaneurol.2018.1836

11. Mecca A et al. Synaptic loss in early Alzheimer’s disease revealed by SV2A PET imaging with [11C]UCB-J. Alzheimer’s Dement. Published online 2020. doi:10.1002/alz.12097

12. Holmes SE, Scheinost D, Finnema SJ, et al. Lower synaptic density is associated with depression severity and network alterations. Nat Commun. 2019;10(1):1529. doi:10.1038/s41467-019-09562-7

13. Corvo C, Mendez-David I, Goutal S, et al. Synaptic Vesicle 2A (SV2A) Positron Emission Tomography (PET) Imaging as a Marker of Therapeutic Response in a Mouse Model of Depression. ACS Pharmacol Transl Sci. 2025;8(2):339–345. doi:10.1021/acsptsci.4c00621

14. Asch RH, Martin NH, Garcia-Milian R, et al. In Vivo Positron Emission Tomography Imaging of Presynaptic Density Reveals Stress-Associated Synaptic Deficits Related to Behavioral and Molecular Alterations in Rats. Biol Psychiatry Cogn Neurosci Neuroimaging. 2026;11(4):463–475. doi:10.1016/j.bpsc.2025.09.021

15. Fries GR, Saldana VA, Finnstein J, Rein T. Molecular pathways of major depressive disorder converge on the synapse. Mol Psychiatry. 2023;28(1):284–297. doi:10.1038/s41380-022-01806-1

16. Vande Casteele T, Laroy M, Van Cauwenberge M, et al. Preliminary evidence for preserved synaptic density in late-life depression. Transl Psychiatry. 2024;14:145. doi:10.1038/s41398-024-02837-8

17. Holmes SE, Finnema SJ, Naganawa M, et al. Imaging the effect of ketamine on synaptic density (SV2A) in the living brain. Mol Psychiatry. 2022;27(4):2273–2281. doi:10.1038/s41380-022-01465-2

18. Laroy M, Vande Casteele T, Van Cauwenberge M, et al. Synaptic density changes following electroconvulsive therapy: A longitudinal pilot study with PET-MR 11C-UCB-J imaging in late-life depression. Brain Stimulat. 2024;17(3):588–590. doi:10.1016/j.brs.2024.04.020

19. Johansen A et al. Escitalopram treatment increases synaptic density over time: a [11C]UCB-J PET study. Mol Psychiatry. Published online 2023. doi:10.1038/s41380-023-02285-8

20. Sheline YI, Wang PW, Gado MH, Csernansky JG, Vannier MW. Hippocampal atrophy in recurrent major depression. Proc Natl Acad Sci U S A. 1996;93(9):3908–3913. doi:10.1073/pnas.93.9.3908

21. Sheline YI, Gado MH, Kraemer HC. Untreated Depression and Hippocampal Volume Loss. Am J Psychiatry. 2003;160:1516–1518. doi:10.1176/appi.ajp.160.8.1516

22. Macqueen G, Campbell S, Mcewen B, et al. Course of Illness, Hippocampal Function, and Hippocampal Volume in Major Depression. Proc Natl Acad Sci U S A. 2003;100:1387–1392. doi:10.1073/pnas.0337481100

23. Videbech P, Ravnkilde B. Hippocampal Volume and Depression: A Meta-Analysis of MRI Studies. Am J Psychiatry. 2004;161:1957–1966. doi:10.1176/appi.ajp.161.11.1957

24. Magariños AM, Verdugo JMG, McEwen BS. Chronic stress alters synaptic terminal structure in hippocampus. Proc Natl Acad Sci U S A. 1997;94(25):14002–14008. doi:10.1073/pnas.94.25.14002

25. Magariños AM, McEwen BS. Stress-induced atrophy of apical dendrites of hippocampal CA3c neurons: comparison of stressors. Neuroscience. 1995;69(1):83–88. doi:10.1016/0306-4522(95)00256-i

26. Radley JJ, Sisti HM, Hao J, et al. Chronic behavioral stress induces apical dendritic reorganization in pyramidal neurons of the medial prefrontal cortex. Neuroscience. 2004;125(1):1–6. doi:10.1016/j.neuroscience.2004.01.006

27. Radley JJ, Rocher AB, Miller M, et al. Repeated Stress Induces Dendritic Spine Loss in the Rat Medial Prefrontal Cortex. Cereb Cortex. 2006;16(3):313–320. doi:10.1093/cercor/bhi104

28. Bennett MR. The prefrontal–limbic network in depression: A core pathology of synapse regression. Prog Neurobiol. 2011;93(4):457–467. doi:10.1016/j.pneurobio.2011.01.001

29. Tjerkaski J, Cervenka S, Farde L, Matheson GJ. Kinfitr — an open-source tool for reproducible PET modelling: validation and evaluation of test-retest reliability. EJNMMI Res. 2020;10(1):77. doi:10.1186/s13550-020-00664-8

30. Nosek BA, Errington TM. What is replication? PLOS Biol. 2020;18(3):e3000691. doi:10.1371/journal.pbio.3000691

31. Crandall CS, Sherman JW. On the scientific superiority of conceptual replications for scientific progress. J Exp Soc Psychol. 2016;66:93–99. doi:10.1016/j.jesp.2015.10.002

32. Holmes SE, Girgenti MJ, Davis MT, et al. Altered metabotropic glutamate receptor 5 markers in PTSD: In vivo and postmortem evidence. Proc Natl Acad Sci U S A. 2017;114(31):8390–8395. doi:10.1073/pnas.1701749114

33. Schoevers RA, van Borkulo CD, Lamers F, et al. Affect fluctuations examined with ecological momentary assessment in patients with current or remitted depression and anxiety disorders. Psychol Med. 51(11):1906–1915. doi:10.1017/S0033291720000689

34. Wirz-Justice A. Diurnal variation of depressive symptoms. Dialogues Clin Neurosci. 2008;10(3):337–343. doi:10.31887/DCNS.2008.10.3/awjustice

35. Raval NR, Johansen A, Donovan LL, et al. A Single Dose of Psilocybin Increases Synaptic Density and Decreases 5-HT2A Receptor Density in the Pig Brain. Int J Mol Sci. 2021;22(2):835. doi:10.3390/ijms22020835

