## Supplement for "Synaptic density (SV2A) tracks illness chronicity rather than symptom severity in major depressive disorder"

This Supplement provides all statistical analyses supporting the main findings. Primary analyses correspond to a priori defined regions of interest, while exploratory analyses include chronicity-related analyses in the primary regions as well as analyses across additional cortical and subcortical regions. Regression models are presented in a stepwise manner for clarity. All p-values are two-sided and corrected for multiple comparisons where appropriate.

**Supplementary Table 1 Primary correlation analyses**
**Supplementary Table 2 Primary regression models**
**Supplementary Table 3 Fully adjusted regression models in primary regions of interest**
**Supplementary Table 4 Exploratory correlation analyses**
**Supplementary Table 5 Exploratory regression models**
**Supplementary Table 6 Sensitivity analyses of current depressive episode duration**
**Supplementary Table 7. Medications tapered before baseline PET**

### Supplementary Table 1. Primary correlation analyses

| ROI | n | r | 95% CI | p | Bonferroni-adjusted p |
|---|---|---|---|---|---|
| ACC | 32 | 0.258 | -0.1 to 0.557 | 0.154 | 0.463 |
| Hippocampus | 32 | 0.26 | -0.097 to 0.558 | 0.15 | 0.450 |
| dlPFC | 32 | 0.177 | -0.183 to 0.495 | 0.332 | >0.99 |

Pearson correlations between depressive symptom severity and SV2A binding (VT) in a priori defined regions of interest. Correlation coefficients (r), 95% confidence intervals, and p-values are reported. Bonferroni-adjusted p-values are shown where indicated.

Abbreviations: ACC, anterior cingulate cortex; dlPFC, dorsolateral prefrontal cortex; SV2A, synaptic vesicle glycoprotein 2A; VT, total distribution volume; CI, confidence interval

### Supplementary Table 2. Primary regression models

| Model | ROI | n | b | 95% CI | p | Bonferroni-adjusted p | $R^2$ | Adjusted $R^2$ |
|---|---|---|---|---|---|---|---|---|
| Model 1: unadjusted | ACC | 32 | 0.153 | -0.061 to 0.367 | 0.154 | 0.463 | 0.066 | 0.035 |
| Model 1: unadjusted | Hippocampus | 32 | 0.114 | -0.043 to 0.27 | 0.15 | 0.450 | 0.068 | 0.037 |
| Model 1: unadjusted | dlPFC | 32 | 0.1 | -0.107 to 0.306 | 0.332 | >0.99 | 0.031 | 0.001 |
| Model 2: age and sex adjusted | ACC | 32 | 0.157 | -0.059 to 0.372 | 0.148 | 0.444 | 0.126 | 0.032 |
| Model 2: age and sex adjusted | Hippocampus | 32 | 0.119 | -0.039 to 0.277 | 0.133 | 0.400 | 0.131 | 0.038 |
| Model 2: age and sex adjusted | dlPFC | 32 | 0.105 | -0.103 to 0.313 | 0.31 | 0.929 | 0.095 | 0.001 |
| Model 3: fully adjusted | ACC | 28 | 0.306 | 0.082 to 0.53 | 0.01 | 0.029 | 0.46 | 0.337 |
| Model 3: fully adjusted | Hippocampus | 28 | 0.231 | 0.077 to 0.385 | 0.005 | 0.015 | 0.509 | 0.397 |
| Model 3: fully adjusted | dlPFC | 28 | 0.255 | 0.04 to 0.47 | 0.022 | 0.067 | 0.432 | 0.303 |

Linear regression models examining the association between depressive symptom severity and SV2A binding (VT) in a priori defined regions of interest. Model 1 represents unadjusted analyses, Model 2 is adjusted for age and sex, and Model 3 is adjusted for age, sex, lifetime number of depressive episodes, and duration of the current depressive episode. Unstandardized regression coefficients (b), 95% confidence intervals, and p-values are reported.

Abbreviations: ACC, anterior cingulate cortex; dlPFC, dorsolateral prefrontal cortex; SV2A, synaptic vesicle glycoprotein 2A; VT, total distribution volume; CI, confidence interval.

### Supplementary Table 3. Fully adjusted regression models in primary regions of interest

| ROI | Predictor | b | 95% CI | p | Bonferroni-adjusted p |
|---|---|---|---|---|---|
| ACC | MADRS total score | 0.306 | 0.082 to 0.530 | 0.010 | 0.029 |
| ACC | Age | -0.060 | -0.172 to 0.052 | 0.279 | 0.837 |
| ACC | Male sex | -1.798 | -4.070 to 0.475 | 0.115 | 0.345 |
| ACC | Lifetime depressive episodes | 0.045 | -0.149 to 0.240 | 0.633 | >0.99 |
| ACC | Duration of current depressive episode | -0.171 | -0.283 to -0.059 | 0.004 | 0.013 |
| Hippocampus | MADRS total score | 0.231 | 0.077 to 0.385 | 0.005 | 0.015 |
| Hippocampus | Age | -0.063 | -0.140 to 0.014 | 0.103 | 0.308 |
| Hippocampus | Male sex | -1.017 | -2.580 to 0.546 | 0.191 | 0.573 |
| Hippocampus | Lifetime depressive episodes | 0.064 | -0.069 to 0.198 | 0.329 | 0.987 |
| Hippocampus | Duration of current depressive episode | -0.121 | -0.198 to -0.044 | 0.004 | 0.011 |
| dlPFC | MADRS total score | 0.255 | 0.040 to 0.470 | 0.022 | 0.067 |
| dlPFC | Age | -0.061 | -0.168 to 0.047 | 0.254 | 0.763 |
| dlPFC | Male sex | -1.435 | -3.619 to 0.748 | 0.187 | 0.560 |
| dlPFC | Lifetime depressive episodes | 0.024 | -0.163 to 0.210 | 0.795 | >0.99 |
| dlPFC | Duration of current depressive episode | -0.164 | -0.272 to -0.057 | 0.004 | 0.013 |

Note. Linear regression models examining associations between depressive symptom severity, clinical covariates, and SV2A binding (VT) in the three a priori defined regions of interest. All models included 28 participants with complete data and were adjusted for age, sex, lifetime number of depressive episodes, and duration of the current depressive episode. Unstandardized regression coefficients (b), 95% confidence intervals, and p-values are reported. Bonferroni-adjusted p-values account for testing across the three primary regions of interest for each predictor. Sex was coded with female as the reference category.

Abbreviations: ACC, anterior cingulate cortex; dlPFC, dorsolateral prefrontal cortex; MADRS, Montgomery-Åsberg Depression Rating Scale; SV2A, synaptic vesicle glycoprotein 2A; VT, total distribution volume; CI, confidence interval.

### Supplementary Table 4. Exploratory correlation analyses

| ROI | n | r | 95% CI | p | FDR-adjusted p |
|---|---|---|---|---|---|
| Amygdala | 32 | 0.34 | -0.01 to 0.616 | 0.057 | 0.296 |
| Bed nucleus of the stria terminalis | 32 | 0.276 | -0.08 to 0.57 | 0.126 | 0.296 |
| Caudate | 32 | 0.176 | -0.184 to 0.494 | 0.336 | 0.367 |
| Centrum semiovale | 32 | 0.238 | -0.121 to 0.542 | 0.19 | 0.325 |
| Frontal cortex | 32 | 0.213 | -0.146 to 0.523 | 0.241 | 0.349 |
| Gray matter | 32 | 0.267 | -0.09 to 0.563 | 0.14 | 0.296 |
| Insula | 32 | 0.306 | -0.048 to 0.592 | 0.088 | 0.296 |
| Putamen | 32 | 0.262 | -0.096 to 0.559 | 0.148 | 0.296 |
| Thalamus | 32 | 0.197 | -0.163 to 0.511 | 0.279 | 0.349 |
| Caudal anterior cingulate cortex | 32 | 0.193 | -0.167 to 0.507 | 0.29 | 0.349 |
| Medial orbitofrontal cortex | 32 | 0.145 | -0.214 to 0.47 | 0.427 | 0.427 |
| Rostral anterior cingulate cortex | 32 | 0.294 | -0.061 to 0.583 | 0.103 | 0.296 |

Pearson correlations between depressive symptom severity and SV2A binding (VT) across exploratory cortical and subcortical regions. Correlation coefficients (r), 95% confidence intervals, and p-values are reported. P-values were corrected for multiple comparisons using the false discovery rate (FDR).

Abbreviations: SV2A, synaptic vesicle glycoprotein 2A; VT, total distribution volume; CI, confidence interval; FDR, false discovery rate.

### Supplementary Table 5. Exploratory regression models

| Model | ROI | n | b | 95% CI | p | FDR-adjusted p | $R^2$ | Adjusted $R^2$ |
|---|---|---|---|---|---|---|---|---|
| Model 1: unadjusted | Amygdala | 32 | 0.219 | -0.007 to 0.445 | 0.057 | 0.296 | 0.116 | 0.086 |
| Model 1: unadjusted | Bed nucleus of the stria terminalis | 32 | 0.055 | -0.016 to 0.127 | 0.126 | 0.296 | 0.076 | 0.045 |
| Model 1: unadjusted | Caudate | 32 | 0.111 | -0.121 to 0.342 | 0.336 | 0.367 | 0.031 | -0.001 |
| Model 1: unadjusted | Centrum semiovale | 32 | 0.028 | -0.014 to 0.069 | 0.19 | 0.325 | 0.057 | 0.025 |
| Model 1: unadjusted | Frontal cortex | 32 | 0.117 | -0.083 to 0.317 | 0.241 | 0.349 | 0.046 | 0.014 |
| Model 1: unadjusted | Gray matter | 32 | 0.132 | -0.046 to 0.31 | 0.14 | 0.296 | 0.071 | 0.04 |
| Model 1: unadjusted | Insula | 32 | 0.167 | -0.027 to 0.361 | 0.088 | 0.296 | 0.094 | 0.064 |
| Model 1: unadjusted | Putamen | 32 | 0.171 | -0.064 to 0.405 | 0.148 | 0.296 | 0.068 | 0.037 |
| Model 1: unadjusted | Thalamus | 32 | 0.082 | -0.07 to 0.233 | 0.279 | 0.349 | 0.039 | 0.007 |
| Model 1: unadjusted | Caudal anterior cingulate cortex | 32 | 0.113 | -0.101 to 0.327 | 0.29 | 0.349 | 0.037 | 0.005 |
| Model 1: unadjusted | Medial orbitofrontal cortex | 32 | 0.088 | -0.135 to 0.311 | 0.427 | 0.427 | 0.021 | -0.011 |
| Model 1: unadjusted | Rostral anterior cingulate cortex | 32 | 0.191 | -0.041 to 0.423 | 0.103 | 0.296 | 0.086 | 0.056 |
| Model 2: age and sex adjusted | Amygdala | 32 | 0.225 | -0.007 to 0.457 | 0.057 | 0.291 | 0.141 | 0.049 |
| Model 2: age and sex adjusted | Bed nucleus of the stria terminalis | 32 | 0.058 | -0.013 to 0.13 | 0.106 | 0.291 | 0.146 | 0.055 |
| Model 2: age and sex adjusted | Caudate | 32 | 0.119 | -0.114 to 0.353 | 0.303 | 0.331 | 0.09 | -0.008 |
| Model 2: age and sex adjusted | Centrum semiovale | 32 | 0.029 | -0.013 to 0.071 | 0.171 | 0.293 | 0.109 | 0.014 |
| Model 2: age and sex adjusted | Frontal cortex | 32 | 0.122 | -0.082 to 0.325 | 0.23 | 0.311 | 0.087 | -0.011 |
| Model 2: age and sex adjusted | Gray matter | 32 | 0.138 | -0.041 to 0.317 | 0.125 | 0.291 | 0.13 | 0.037 |
| Model 2: age and sex adjusted | Insula | 32 | 0.172 | -0.021 to 0.365 | 0.079 | 0.291 | 0.169 | 0.08 |
| Model 2: age and sex adjusted | Putamen | 32 | 0.177 | -0.065 to 0.419 | 0.145 | 0.291 | 0.087 | -0.011 |
| Model 2: age and sex adjusted | Thalamus | 32 | 0.089 | -0.062 to 0.239 | 0.237 | 0.311 | 0.121 | 0.027 |
| Model 2: age and sex adjusted | Caudal anterior cingulate cortex | 32 | 0.119 | -0.093 to 0.331 | 0.259 | 0.311 | 0.131 | 0.038 |
| Model 2: age and sex adjusted | Medial orbitofrontal cortex | 32 | 0.092 | -0.133 to 0.317 | 0.408 | 0.408 | 0.08 | -0.018 |
| Model 2: age and sex adjusted | Rostral anterior cingulate cortex | 32 | 0.192 | -0.045 to 0.429 | 0.108 | 0.291 | 0.12 | 0.025 |
| Model 3: fully adjusted | Amygdala | 28 | 0.393 | 0.155 to 0.631 | 0.002 | 0.015 | 0.461 | 0.339 |
| Model 3: fully adjusted | Bed nucleus of the stria terminalis | 28 | 0.105 | 0.029 to 0.181 | 0.009 | 0.023 | 0.428 | 0.298 |
| Model 3: fully adjusted | Caudate | 28 | 0.26 | 0.019 to 0.502 | 0.036 | 0.039 | 0.428 | 0.298 |
| Model 3: fully adjusted | Centrum semiovale | 28 | 0.051 | 0.007 to 0.096 | 0.026 | 0.031 | 0.373 | 0.231 |
| Model 3: fully adjusted | Frontal cortex | 28 | 0.266 | 0.057 to 0.475 | 0.015 | 0.026 | 0.434 | 0.305 |
| Model 3: fully adjusted | Gray matter | 28 | 0.257 | 0.066 to 0.448 | 0.011 | 0.023 | 0.423 | 0.292 |
| Model 3: fully adjusted | Insula | 28 | 0.317 | 0.13 to 0.505 | 0.002 | 0.015 | 0.544 | 0.441 |

| Model | ROI | n | b | 95% CI | p | FDR-adjusted p | $R^2$ | Adjusted $R^2$ |
|---|---|---|---|---|---|---|---|---|
| Model 3: fully adjusted | Putamen | 28 | 0.332 | 0.082 to 0.581 | 0.011 | 0.023 | 0.419 | 0.287 |
| Model 3: fully adjusted | Thalamus | 28 | 0.169 | -0.001 to 0.339 | 0.051 | 0.051 | 0.329 | 0.177 |
| Model 3: fully adjusted | Caudal anterior cingulate cortex | 28 | 0.257 | 0.035 to 0.479 | 0.025 | 0.031 | 0.441 | 0.314 |
| Model 3: fully adjusted | Medial orbitofrontal cortex | 28 | 0.257 | 0.035 to 0.479 | 0.025 | 0.031 | 0.489 | 0.373 |
| Model 3: fully adjusted | Rostral anterior cingulate cortex | 28 | 0.347 | 0.097 to 0.597 | 0.009 | 0.023 | 0.442 | 0.315 |

Linear regression models examining the association between depressive symptom severity and SV2A binding (VT) across exploratory cortical and subcortical regions. Model 1 represents unadjusted analyses, Model 2 is adjusted for age and sex, and Model 3 is adjusted for age, sex, lifetime number of depressive episodes, and duration of the current depressive episode. Unstandardized regression coefficients (b), 95% confidence intervals, and p-values are reported. FDR-adjusted p-values account for multiple comparisons across exploratory regions within each model.

Abbreviations: SV2A, synaptic vesicle glycoprotein 2A; VT, total distribution volume; CI, confidence interval; FDR, false discovery rate.

### Supplementary Table 6. Sensitivity analyses of the association between current depressive episode duration and SV2A binding in primary regions

| ROI | Pearson r (p) | Spearman ρ (p) | Log-duration b, Model 3 (p) | Leave-one-out b range | Negative b | $p < 0.05$ |
|---|---|---|---|---|---|---|
| ACC | −0.366 (0.043) | −0.345 (0.057) | −2.195 (0.003) | −0.248 to −0.149 | 28/28 | 28/28 |
| Hippocampus | −0.361 (0.046) | −0.372 (0.039) | −1.400 (0.007) | −0.139 to −0.109 | 28/28 | 28/28 |
| dlPFC | −0.369 (0.041) | −0.340 (0.061) | −2.055 (0.004) | −0.223 to −0.141 | 28/28 | 28/28 |

**Note.** Pearson and Spearman correlations assess the bivariate association between current depressive episode duration and SV2A binding. Log-duration coefficients are from fully adjusted Model 3 including MADRS total score, age, sex, lifetime depressive episodes, and log-transformed current episode duration. Raw and log-transformed regression coefficients are not directly comparable because the predictor scales differ. Influence diagnostics identified potentially influential observations in the fully adjusted models; however, the episode-duration coefficient remained negative and statistically significant in every leave-one-out iteration across all three primary regions.

**Abbreviations:** ACC, anterior cingulate cortex; dlPFC, dorsolateral prefrontal cortex; SV2A, synaptic vesicle glycoprotein 2A.

### Supplementary Table 7. Medications tapered before baseline PET

| Medication tapered | Dose |
|---|---|
| Clomipramine; Lisdexamfetamine | 75 mg; 70 mg* |
| Vortioxetine | 5 mg |
| Vortioxetine | 15 mg |
| Mirtazapine | 30 mg |
| Quetiapine | Dose unknown |
| Sertraline | 100 mg |
| Sertraline | 50 mg |

**Note**. Seven participants tapered medication before baseline assessment. *One participant tapered two medications.
